# A Streamlined CyTOF Workflow To Facilitate Standardized Multi-Site Immune Profiling of COVID-19 Patients

**DOI:** 10.1101/2020.06.26.20141341

**Authors:** Daniel Geanon, Brian Lee, Geoffrey Kelly, Diana Handler, Bhaskar Upadhyaya, John Leech, Manon Herbinet, Diane Del Valle, Sacha Gnjatic, Seunghee Kim-Schulze, Miriam Merad, Adeeb H. Rahman

## Abstract

Mass cytometry (CyTOF) represents one of the most powerful tools in immune phenotyping, allowing high throughput quantification of over 40 single parameters at single-cell resolution. However, wide deployment of CyTOF-based immune phenotyping studies are limited by complex experimental workflows and the need for specialized CyTOF equipment and technical expertise. Furthermore, differences in cell isolation and enrichment protocols, antibody reagent preparation, sample staining and data acquisition protocols can all introduce technical variation that can potentially confound integrative analyses of large data-sets of samples processed across multiple labs.

Here, we describe the validation of a streamlined whole blood CyTOF workflow that addresses many of these challenges and facilitates wider adoption of CyTOF immune monitoring across sites with limited technical expertise or sample-processing resources or equipment. Our workflow utilizes commercially available reagents including the Fluidigm MaxPar Direct Immune Profiling Assay, a dry tube 30-marker immunophenotyping panel, and SmartTube Proteomic Stabilizer, which allows for simple and reliable fixation and cryopreservation of whole blood samples. We validate a workflow that allows for streamlined staining of whole blood samples with minimal processing requirements or expertise at the site of sample collection, followed by shipment to a central CyTOF core facility for batched downstream processing and data acquisition. We further demonstrate the application of this workflow to characterize immune responses in a cohort of hospitalized COVID-19 patients, highlighting key disease-associated changes in immune cell frequency and phenotype.

## Introduction

High dimensional cytometry is a powerful approach to characterize the immune system to identify immunological mechanisms and correlates of disease progression. Mass cytometry allows for an evaluation of over 40 parameters in a single sample, thereby enabling comprehensive characterization of immune cells in limited samples. However, the need for large numbers of reagents, long sample processing workflows and expensive and complicated CyTOF hardware present challenges that limit wide adoption of mass cytometry assays. These challenges are further amplified in studies involving clinical sample collection at multiple sites that may differ in their available sample processing resources and levels of technical expertise.

To address these challenges, we have optimized a standardized, streamlined sample processing workflow to allow whole blood to be easily stained, stabilized, preserved and transferred to a central CyTOF core for final processing and data acquisition. This whole blood sample processing workflow leverages the commercially available Fluidigm MaxPar Direct Immune Profiling Assay (MDIPA), which incorporates a dry tube containing a 30-marker broad immunophenotyping panel. Pre-mixed dry antibody panels offer significant advantages over a conventional liquid antibody for flow cytometry workflows by significantly reducing the processing time, technical variation and potential errors associated with pipetting multiple individual antibodies, and these advantages are amplified when using larger antibody panels^1^. Applying this assay to whole blood rather than PBMCs offers further sample sparing advantages by allowing the assay to be performed with only 270µL of whole blood, eliminating the labor and technical variation associated with PBMC isolation, and allowing analysis of granulocyte subsets that are otherwise removed by density centrifugation. The MDIPA workflow has previously been found to show excellent intra- and inter-site reproducibility using both whole blood and PBMCs^2^. Despite these advantages, the standard MDIPA protocol as provided by Fluidigm still requires approximately 2 hours of upfront sample processing time including several centrifugation steps, followed by an overnight incubation and suggests data acquisition within 48 hours of sample staining. This workflow generally limits sample collection to labs that have the necessary technical expertise and resources for sample processing and on-site mass cytometry instrumentation.

To facilitate broader adoption of this assay in studies involving multiple clinical sample collection sites we have adapted this dry MDIPA antibody panel as part of a protocol utilizing commercially available SmartTube proteomic stabilizer to significantly reduce sample processing time and hardware requirements at the site of sample collection (Figure 1). By minimizing initial sample processing requirements and transferring downstream processing steps to a central core, this workflow facilitates robust, highly standardized CyTOF immune monitoring in studies involving multiple clinical sites that cannot accommodate complex sample processing workflows and that do not have their own mass cytometry instrumentation. We further demonstrate the application of this workflow to study blood samples collected from COVID-19 patients, highlighting its utility in facilitating large scale standardized immune monitoring initiatives.

**Figure 1.**
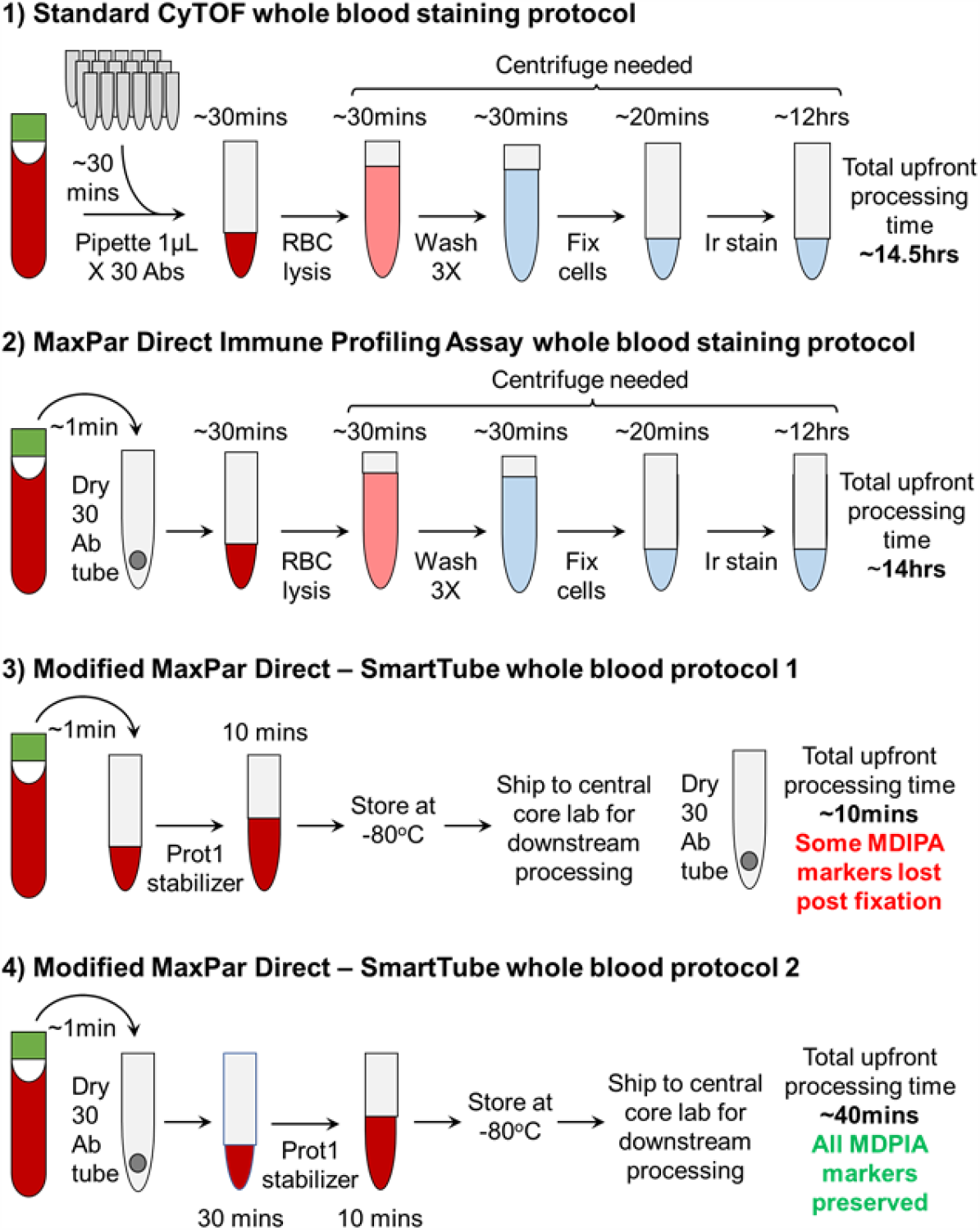
Graphical overview of the MDIPA-SmartTube workflow.

## Methods

### Samples

Samples were collected from consented healthy donors and hospitalized COVID-19 patients under protocols approved by the Mount Sinai Institutional Review Board. Fresh whole blood samples were collected in either heparin, EDTA, or sodium citrate CPT tubes (BD Biosciences, Franklin Lakes, NJ, USA) as indicated for specific experiments.

### Whole blood staining

Whole blood samples were stained by adding 270µL blood directly to a MDIPA tube containing the lyophilized antibody panel (Table 1), mixing and incubating for 30 minutes at room temperature. For the conventional MDIPA workflow, stained blood samples were fixed and red blood cells were lysed using Cal-lyse buffer (Thermo Fisher Scientific, Waltham, MA, USA) as per Fluidigm’s recommended protocols. The samples were washed with Cell Staining Buffer (Fluidigm, San Francisco, CA, USA), fixed and permeabilized using Fix Perm buffer (Fluidigm), and stained with 125nM Iridium DNA intercalator.

**Table 1.**
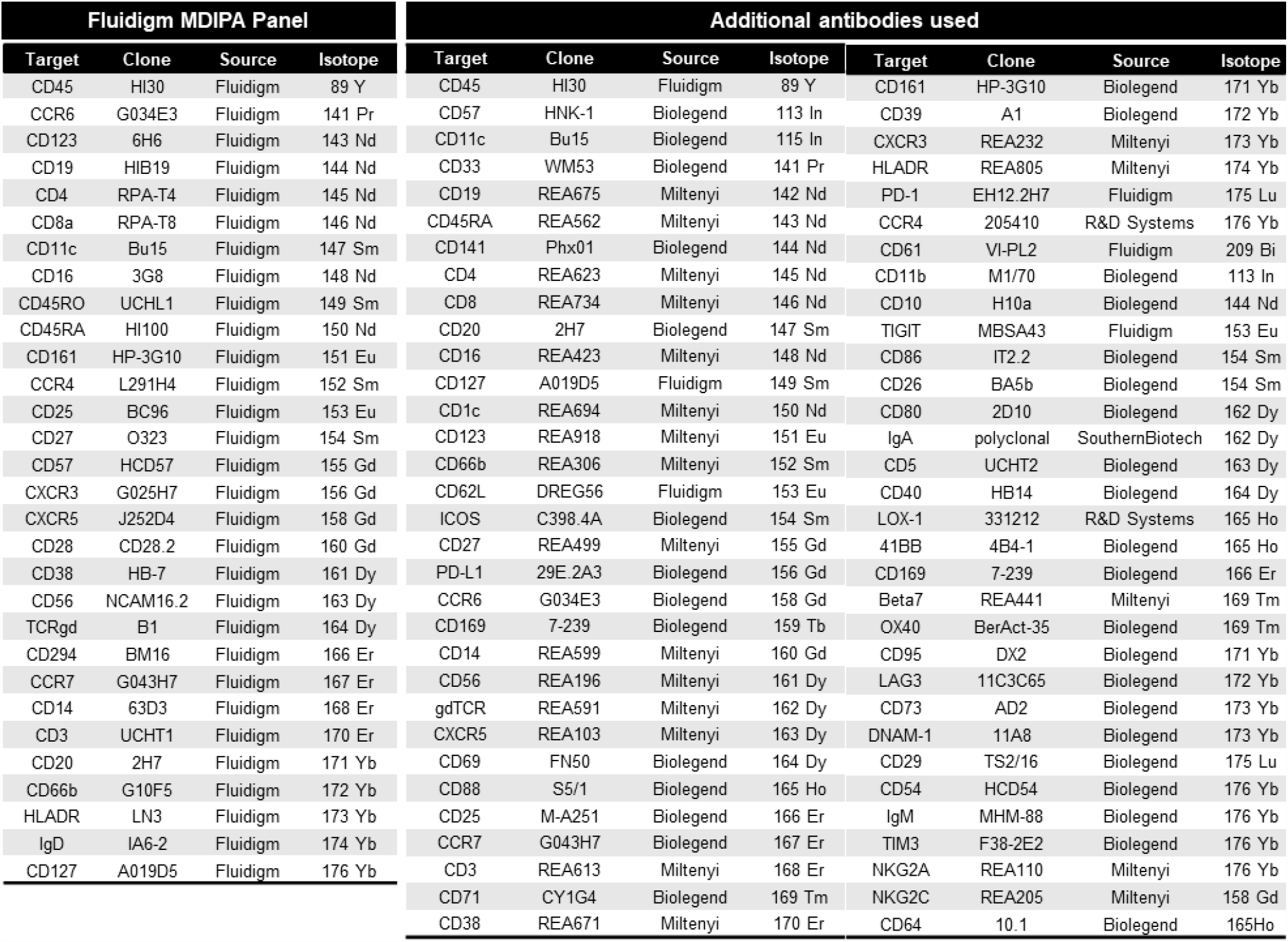
Antibodies

For our alternative protocol, 300uL of the MDIPA-stained blood was fixed and stabilized by addition of 420µL of Prot1 stabilizer (SmartTube Inc. San Carlos, CA, USA). After incubation for 10mins at room temperature the samples were transferred to labeled cryovials and immediately transferred to −80°C for long term storage and/or shipment. Samples were subsequently thawed using the SmartTube Prot 1 Thaw/Erythrocyte Lysis protocol according to the manufacturer’s instructions. After subsequent washing in PBS + 0.2% BSA, samples were simultaneously fixed with 2.4% PFA in PBS with 0.08% saponin and 125nM Iridium intercalator for 30 minutes at room temperature, after which samples were washed and stored in PBS + 0.2% BSA until acquisition. To facilitate data acquisition and doublet removal, multiple samples were also barcoded using Fluidigm Pd barcoding kits and then washed and pooled for data acquisition. If stained samples could not be acquired within 48hrs of staining completion, we found that storage of the samples at −80°C was optimal to preserve staining quality^3^.

In experiments to evaluate the performance of additional antibody clones (Table 1) pre- and post-Prot1 fixation, replicate aliquots of the same blood samples were either stained as fresh whole blood and then fixed and frozen or were fixed and frozen as unstained aliquots and then thawed and lysed as described above. The thawed samples were then resuspended in a volume of Cell Staining Buffer equal to the volume of the original blood aliquot for staining. SmartTube fixation and thaw/lyse results in partial permeabilization of cells and, consistent with prior results^4^, we found that addition of 100U/ml of heparin was critical to prevent non-specific staining of eosinophils when adding antibodies to SmartTube-fixed whole blood. However, this was not found to be necessary when staining fresh whole blood.

It is important to note that the standard SmartTube thawing protocol as per the manufacturer’s instructions worked well for all healthy donor samples used in our initial validation experiments; however, when applying this protocol to blood collected from hospitalized COVID-19 patients we observed several instances in which the stabilized samples appeared to be partially clotted and exhibited high amounts of debris after thawing and lysis, which we suspect may be related to polymerized fibrin or other plasma factors related to COVID-19 disease-associated coagulopathy. If not addressed, this debris contributed to overall poor sample and staining quality and in some cases precluded analysis of samples. We found that following the red blood cell lysis washes with 3 additional large volume washes using ∼10mL of PBS + 0.2% BSA with centrifugation at 250 rcf and followed by filtration through a 70 micron filter depleted the majority of this debris and permitted effective analysis of blood samples that would otherwise have been discarded.

### Data Acquisition and Processing

Immediately prior to data acquisition, samples were then washed with Cell Staining buffer and Cell Acquisition Solution (Fluidigm) and resuspended at a concentration of 1 million cells per ml in Cell Acquisition Solution containing a 1:20 dilution of EQ Normalization beads (Fluidigm). The samples were then acquired on a Helios Mass Cytometer equipped with a wide-bore sample injector at an event rate of <400 events per second. After acquisition, repeat acquisitions of the same sample concatenated and normalized using the Fluidigm software, and barcoded samples were de-multiplexed using the Zunder single cell debarcoder^5^.

### Data analysis

Debarcoded files were uploaded to Cytobank for analysis. Immune cells were identified based on Ir-193 DNA intensity and CD45 expression; Ce140+ normalization beads, CD45-low/Ir-193-low debris and cross-sample and Gaussian ion-cloud multiplets were excluded from subsequent downstream analysis. Major immune cell types were defined by manual gating (supplemental Fig. 1), and the cell frequencies and median marker intensities of each subset were exported for downstream statistical analyses and visualized as heatmaps using Clustergrammer^6^. In some cases, immune cell populations were also identified using automated approaches including MaxPar PathSetter and Astrolabe, the result of which largely correlated well with our manual gating approaches. However, we found that manual gating was most effective in accurately identifying specific cell populations in the presence of variable marker intensity, as in the case of our fixation experiments.

The impact of each tested condition on relative staining quality was evaluated in two ways: 1) overall correlations were determined by calculating the Pearson’s correlation coefficients for the median expression of each marker across each defined immune subset; and 2) a staining index was calculated using defined populations showing the highest and lowest expression levels of each marker: SI = (Median_pos_ - Median_neg_) / 2 X Std.Dev_neg_.

When applying the MDIPA panel to larger numbers of whole blood samples, we also observed some instances of aberrant CD19 staining on non-B cells, which occurred to a varying degree across subjects but was particularly notable in some samples (approximately 10% of subjects). (Supplemental Figure 2A-B). The phenomenon was observed using both the conventional MDIPA Cal-lyse and SmartTube-based fixation protocols, and could not be attributed to previously described mass cytometry artifacts such as cell-cell multiplets, isotopic spillover or oxidation^7^, or mass cytometer instrument configuration^8^. Instead, we determined that this artifact was caused by donor-specific serum factors interacting with specific reagents in specific lots of the MDIPA (data not shown). While these artifacts could negatively impact unbiased clustering approaches, they could easily be overcome in manual gating analyses by avoiding CD19 as an exclusion parameter for non-B cells and instead using CD20, which did not show any evidence of artifactual staining (Supplemental Figure 2C-D).

## Results

### SmartTube stabilization prior to MDIPA staining negatively impacts several antibodies

SmartTube proteomic stabilizer allows whole blood to be fixed and preserved with the addition of a single addition of buffer followed by a 10 minute incubation, after which blood can be transferred to long-term storage at −80°C and shipped to a remote core for downstream sample staining and processing. This workflow offers the advantage of a very rapid workflow that entails minimal sample processing at the site of collection, and has previously been effectively used to facilitate complex immune monitoring studies^9,10^. However, a limitation of this workflow is that the whole blood stabilization entails fixation, which is expected to impact some antibody epitopes. We evaluated this by taking two parallel aliquots of whole blood, processing one with the conventional MDIPA workflow, and fixing and freezing the second with SmartTube Prot1 stabilizer, followed by staining with the MDIPA panel. The data were analyzed by manual gating all major immune cell subsets and evaluating expression patterns of all 30 markers across all the gated populations (Figure 2A). We found that staining of some markers was preserved post-fixation, while others were significantly compromised in comparison to the conventional MDIPA workflow as shown by a significant reduction in the correlation between marker expression across populations (Figure 2B). Notably, most chemokine receptors were dramatically affected, in many cases showing high non-specific background staining resulting in an inability to distinguish true positive and negative populations, and an overall loss of staining index (Figure 2C). These results are consistent with our prior observations of the effect of formaldehyde fixation on chemokine receptor expression^11^. We also observed a reduced staining index for CD25 and CD127, two markers used to resolve CD4+ T regulatory cells. Thus, while immediate SmartTube fixation offers advantages in rapid blood processing, it is not compatible with all the antibodies used in the MDIPA panel.

**Figure 2.**
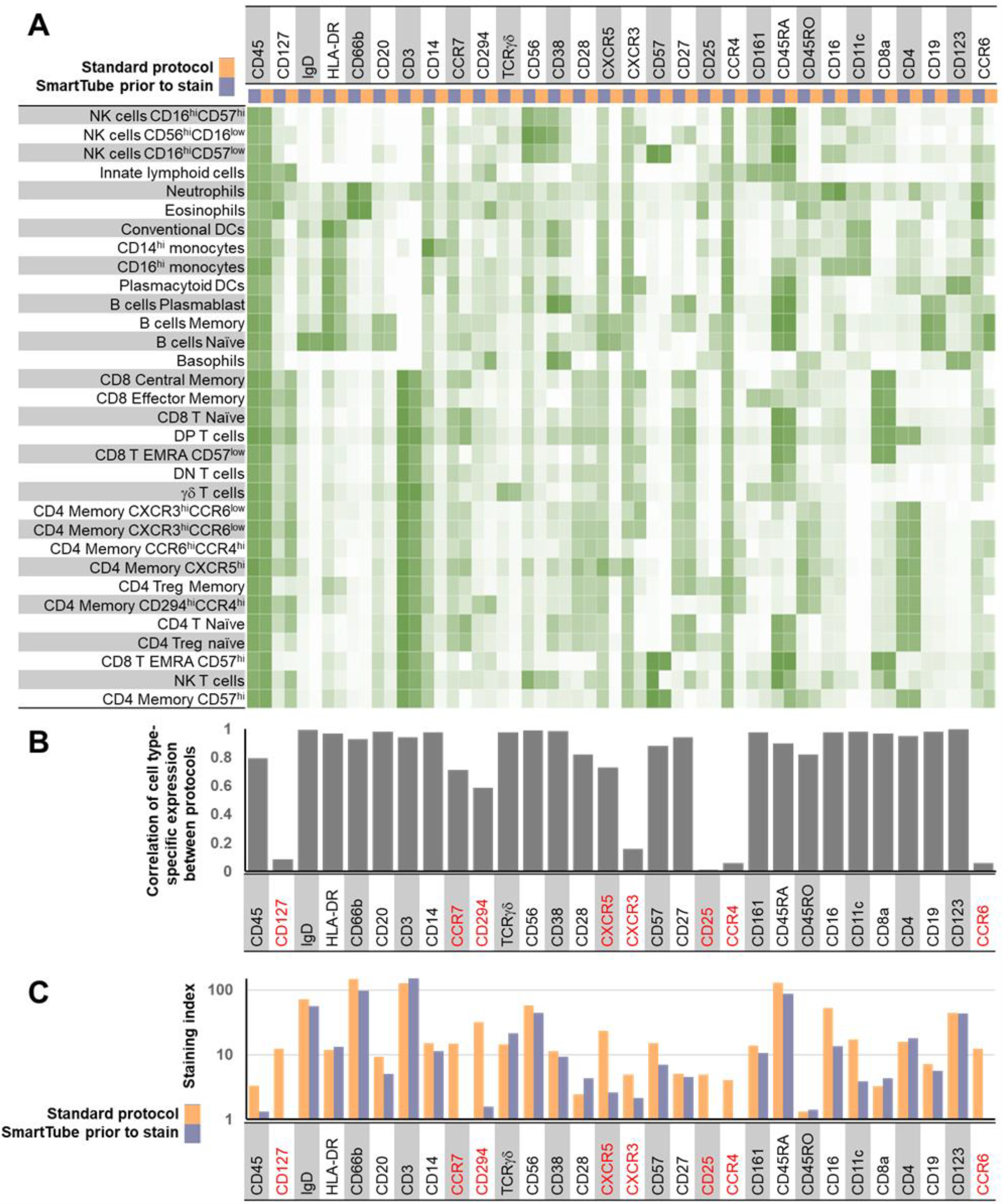
Application of the MDIPA panel to whole blood post-SmartTube fixation negatively impacts resolution of some markers in the panel. Parallel aliquots of whole blood from the same heparin collection tube were stained using the conventional MDIPA workflow or post-SmartTube fixation. (A) Heatmap of median marker expression of each marker on manually gated immune subsets using the two workflows. (B) Pearson’s coefficient of the correlation of the expression of each marker across all subsets using the two workflows. (C) Staining index of each marker using the two workflows.

### SmartTube stabilization following staining faithfully preserves all antibody expression patterns and results in comparable cell frequencies to the conventional MDIPA workflow

We next evaluated a workflow where whole blood was first stained with the MDIPA panel and then fixed and frozen with the SmartTube stabilizer, which entails less than 45 minutes of upfront sample processing time with no centrifugation requirements, after which samples can be frozen and shipped to a remote site for subsequent processing. The workflow entails the addition of the relatively small volume of SmartTube stabilizer (1.4 fold dilution of the starting blood volume). The fact that the cells were being fixed without much dilution of the staining antibodies raised the potential concern of elevated staining background and reduced staining quality due to cross linking of free antibodies to the cells. To evaluate this, we stained parallel aliquots of blood from 3 donors using the MDIPA antibody panel, after which one aliquot was processed according to the standard MDIPA protocol, while the other was fixed, frozen with SmartTube Prot1 and subsequently processed following the SmartTube thaw/lyse protocol. The data were manually gated and markers were compared across populations as in Figure 1. When evaluating marker expression between these two protocols, we found that the SmartTube workflow resulted in nearly identical staining patterns to the conventional MDIPA workflow (Figure 3). Almost all markers showed correlations of over 98% between the two protocols, and in many cases staining indexes were slightly higher with the SmartTube workflow indicating better resolution of cell populations (Figure 3B-C). In addition, we found that the SmartTube-based workflow resulted in an approximately 35% greater recovery of CD45+ cells from the same starting volume of blood (Figure 4A). Overall relative cell population frequencies were highly correlated between both protocols (Figure 4B), and both protocols were able to clearly show consistent inter-individual differences in the frequency of CD4 T cell memory subsets defined by differential chemokine receptor staining (Figure 4C).

**Figure 3.**
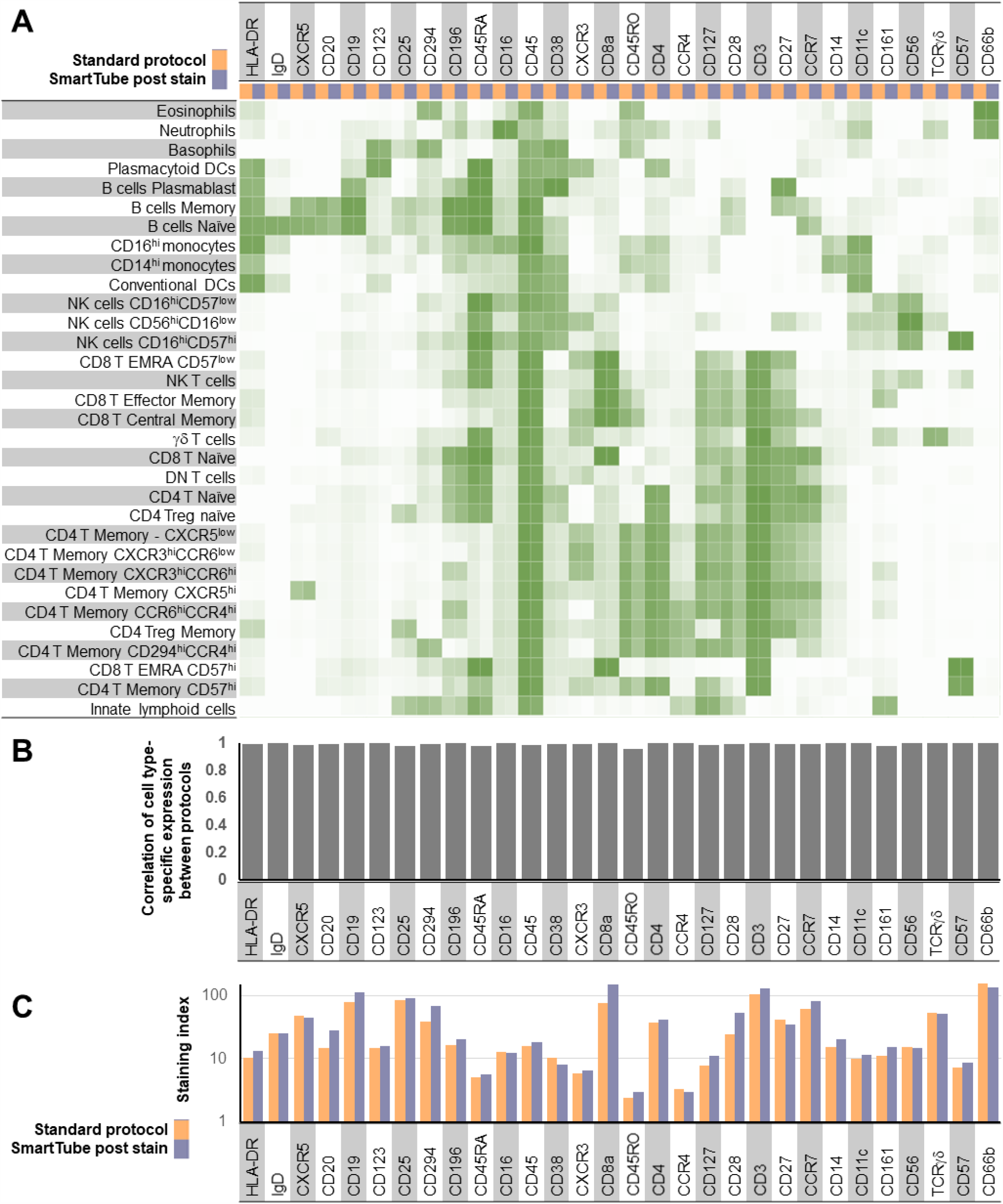
SmartTube-fixation of MDIPA-stained blood accurately reproduces staining patterns obtained with the conventional MDIPA workflow. Parallel aliquots of whole blood from the same heparin collection tube were stained using MDIPA panel and subsequently processed using the conventional MDIPA workflow or the modified SmartTube workflow. (A) Heatmap of median marker expression of each marker on manually gated immune subsets using the two workflows. (B) Pearson’s coefficient of the correlation of the expression of each marker across all subsets using the two workflows. (C) Staining index of each marker using the two workflows.

**Figure 4.**
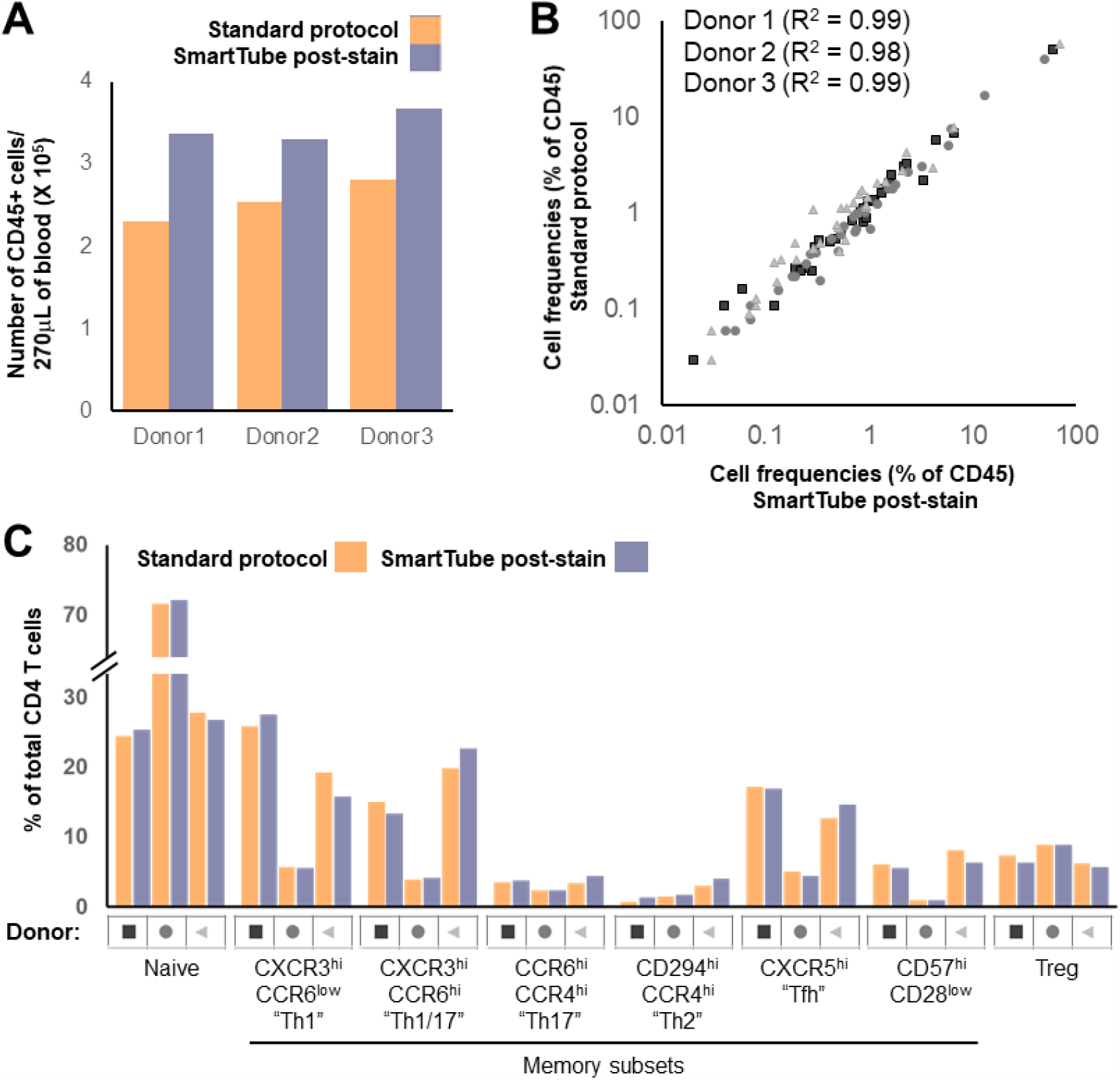
SmartTube-fixation of MDIPA-stained blood improves recovery and accurately reproduces cell frequencies obtained with the conventional MDIPA workflow. Parallel aliquots of whole blood from 3 donors were stained and analyzed as in Figure 3. (A) Overall recovery of CD45+ cells using the two workflows. (B) Correlation of overall cell frequencies for each gated cell type across all three donors. (C) Frequency of CD4+ memory T helper subsets defined by chemokine receptor expression using the two workflows.

### The SmartTube-based workflow can be applied to whole blood collected in multiple tube types

Accurate immunophenotyping requires active steps to identify and minimize potential sources of non-specific antibody staining. One well-known source of non-specific antibody staining is binding by Fc-receptors. While this can be mitigated using Fc-blocking reagents, this is generally not needed in whole blood staining workflows since endogenous serum antibodies effectively occupy and block Fc-receptors. Another source of non-specific antibody staining that is more specific to CyTOF whole blood workflows is a charge-based interaction between cationic granule proteins which can result in non-specific antibody binding by eosinophils. This non-specific interaction can be effectively blocked using heparin, a strongly anionic compound^4^. While this problem primarily presents when performing intracellular staining on fixed whole blood, we were concerned that SmartTube fixation of the blood with relatively minimal antibody dilution may result in non-specific binding of MDIPA antibodies. The majority of our initial tests were conducted using blood collected in sodium heparin vacutainers, which would be expected to prevent any such issues; however, we wanted to evaluate whether higher levels of non-specific eosinophil antibody binding would occur when staining non-heparinized blood and, if so, whether heparin supplementation could mitigate such artefacts. To evaluate this, we collected and stained three aliquots of blood collected from the same individual in either a sodium heparin tube, an EDTA tube, or a sodium citrate CPT tube. Samples were stained with the MDIPA panel and fixed with SmartTube stabilizer and analyzed as described above. We found that overall staining quality was very similar between the tube types, with no evidence of elevated eosinophil background staining in EDTA and citrate CPT tubes relative to heparin tubes (Supplemental Figure 2A). Cell-type specific marker expression patterns (Supplemental Figure 2B-C) and frequencies (Supplemental Figure S2D) were highly correlated between the blood samples collected in all three tube types. However, we did observe some changes in relative staining intensity for some markers, most notably CD8a, which showed an almost 10-fold reduction in staining intensity in blood collected in EDTA tubes relative to that collected heparin tubes (Supplemental Figure S2C). Furthermore, supplementing additional heparin into the EDTA tubes did not have any measurable impact on staining quality (Supplemental Figure 3). Overall, these data show that while staining quality is optimal in heparin tubes, overall staining patterns and relative population frequencies (data not shown) are largely preserved across tube types, indicating that this SmartTube stabilization workflow is broadly applicable to perform CyTOF immunophenotyping on blood samples collected in multiple tube types.

In addition to collection tube type, we also considered delays in the time between sample collection and staining as another factor that could potentially impact immunophenotyping results. While fresh whole blood samples should ideally be processed immediately, this may pose practical challenges in some circumstances. To evaluate the potential impact that delayed processing may have on immune cell frequencies and phenotype, aliquots of blood collected in a CPT tube were stained with the MDIPA panel immediately upon collection, or after storage for 4hrs and 9hrs on a bench at room temperature. The samples were fixed with SmartTube stabilizer and analyzed as described above. Overall immune cellular phenotype and marker expression patterns were largely preserved across both time points relative to baseline (Supplemental Fig. 4). We did observe a relative reduction in the frequency of neutrophils and eosinophils and a corresponding increase in mononuclear cell frequencies which was apparent after even 4hrs of storage. However, the frequencies of more granular immune subsets as a proportion of total mononuclear cells was largely preserved at 4hrs, with all relative frequencies remaining within ∼10% of those found in the baseline samples. Greater deviations were seen by 9hrs, with an apparent increase in the relative frequencies of plasmablasts, dendritic cells and monocytes, and a corresponding decrease in the relative frequency of T cell subsets. Together, these results suggest that reliable immunophenotyping results can be expected from blood samples processed within 4hrs of collection.

### Whole blood stained with the core 30-marker MDIPA can be stained with additional fixation-resistant antibodies at a central processing site to facilitate more in-depth analyses of specific subsets

The 30 marker MDIPA panel offers a consistent set of core markers to identify almost all major circulating immune cell types and allows for robust and highly standardized immune profiling. However, the current range of available metal isotopes allows for supplementation of this core panel with at least 16 additional antibodies to facilitate deeper characterization of specific cell types of interest. While additional markers can certainly be added to the whole blood at the time of MDIPA staining, this presents an additional need for reagents at the collection site and adds complication to the highly streamlined workflow of mixing blood with a single lyophilized reagent. As an alternative, additional markers can instead be added to the MDIPA-stained blood as part of downstream sample processing at the central lab. However, as described in Figure 1, a caveat of staining blood post-SmartTube fixation is that some antibody epitopes are compromised by this fixation process, so supplemental add-ins must be limited to epitopes that are preserved post fixation. Prior work has already identified multiple antibody epitopes that are sensitive and resistant to formaldehyde fixation, and we further validated and expanded upon these results by systematically testing the performance of several relevant immune profiling antibodies on aliquots of the same blood sample pre- and post-SmartTube fixation. To better evaluate the impact of fixation on some dynamic activation markers (e.g., CD86 and PD-L1), we first stimulated whole blood with Phytohaemagglutinin (PHA) to induce marker upregulation. Fresh and stimulated whole blood samples were then stained with a set of phenotyping markers, after which parallel aliquots were stained with a range of additional markers either prior to or after Prot1 stabilization. Major immune cell subsets were defined based on the core phenotyping markers, and cell-type specific marker expression patterns for each of the tested antibodies was evaluated on the paired pre- and post-fixation. While the majority of antibodies showed highly correlated expression patterns, some showed extremely low correlations indicating a severe impact of fixation (Figure 5A). In most cases, the low correlations reflected a loss of detectable expression post-fixation which was the case for NKG2A, NKG2C, CD88, and CD40; however, in the case of IgA and IgM, expression was only detectable on the post-fixed samples (Figure 5B and C). This is likely due to interference from soluble serum IgA and IgM antibodies in fresh whole blood, which are effectively washed away during the Prot1 fixation and lysing protocol. Even amongst antibodies that showed highly correlated expression patterns between the pre- and post-fixed samples, we observed some that showed a reduced staining index post fixation (e.g., CD71) and others that showed increased staining index (e.g., CD29). However, some of these differences could be minimized by re-titration of the antibodies on the fixed blood samples, and all of these antibodies ultimately represent reasonable potential candidates to supplement MDIPA-stained blood samples at the site of central downstream processing.

**Figure 5.**
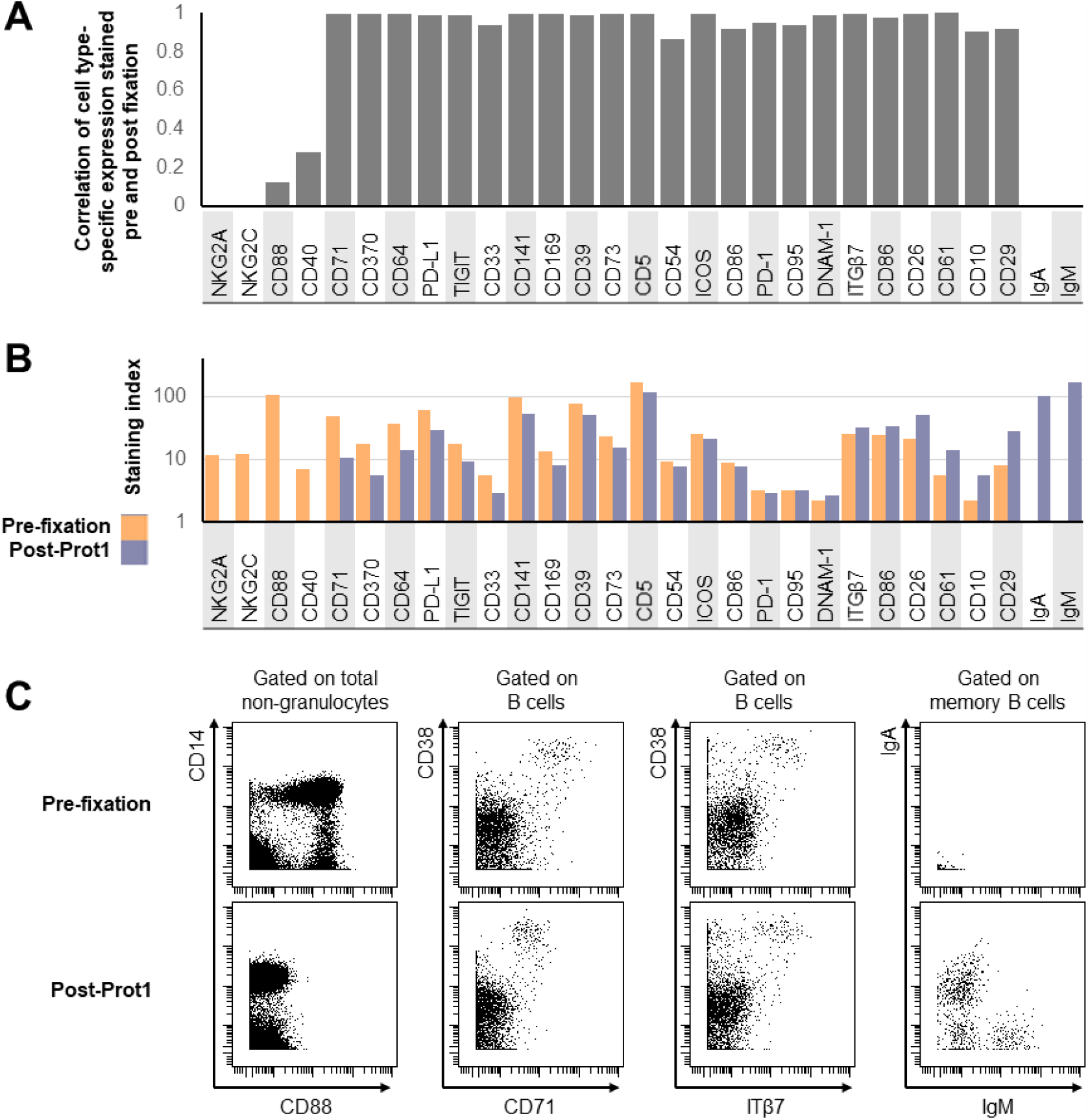
Evaluation of fixation-stable markers to supplement the core MDIPA panel on SmartTube-fixed whole blood. Aliquots of whole blood were stained with several panels of antibodies either prior to or following SmartTube fixation to evaluate the fixation sensitivity of different antibody clones. Each panel shared a core set of fixation-stable markers, which were used to gate major immune cell subsets. (A) Pearson’s coefficient of the correlation in the staining pattern of each antibody across the defined immune cell subsets when stained on fresh blood or post-SmartTube fixation. (B) Staining index of each antibody when stained on fresh blood or post-SmartTube fixation. (C) Representative biaxial plots highlighting staining patterns for antibodies where SmartTube fixation resulted in complete signal loss (CD88), reduced but still resolvable signal (CD71) or improved signal (ITß7, IgM and IgA).

### Application of the MDIPA-SmartTube workflow to characterize immune dysregulation in COVID-19 patients

Our optimized MDIPA-SmartTube protocol offers a highly standardized and streamlined workflow to perform comprehensive immune monitoring of whole blood samples. This approach is particularly well suited to large-scale and multi-center studies where blood samples are being processed under BSL2+/3 conditions by non-specialist technicians. The minimal blood volumes required also allow for a relatively rapid comprehensive assessment of circulating immune cell composition and phenotype, while still allowing for the majority of collected blood to be used for PBMC isolation for biobanking efforts. This sample-sparing aspect is also useful in settings where limited blood collection volumes, or underlying clinical conditions such as lymphopenia, may limit the number of PBMCs that can be successfully isolated and banked for immunophenotyping studies. Furthermore, the final concentration of formaldehyde in SmartTube Prot1 fixative is >2.5%, which is sufficient to completely neutralize viral pathogens and allow subsequent processing of the blood to be safely performed under less stringent BSL conditions.

These advantages make this workflow make it particularly well suited to standardized large-scale immune monitoring of COVID-19 patients. We have already integrated this workflow as part of a large centralized COVID-19 sample collection and biobanking effort at the Mount Sinai Health System, and it is also being deployed as part of the NIAID Immunophenotyping assessment in a COVID-19 Cohort (IMPACC) study, which aims to perform standardized longitudinal immune monitoring of 2000 patients from 12 clinical collection sites across the United States. While the results of these larger studies are pending, here we provide a proof-of-principle demonstration of the power of this assay in resolving changes in circulating immune cell frequency and phenotype associated with COVID-19 disease.

Whole blood samples were collected from 8 healthy donors and 18 hospitalized patients with PCR-confirmed cases of COVID-19 disease within the first 3 days of admission. Aliquots of blood from CPT-Citrate tubes were stained with the MDIPA panel and then stabilized and banked with Prot1 stabilizer as described above. The stabilized blood samples were then thawed, barcoded using a combinatorial Pd-barcoding system and then pooled and stained with a panel of fixation-resistant antibodies. This barcoding approach offers the additional advantage of further streamlining downstream sample processing and minimizing any technical variability in subsequent staining acquisition so as to maximize the ability to accurately resolve subtle differences in protein expression. As discussed in the methods, we found that blood samples from some COVID-19 patients exhibit high amount of debris, which we believe may be polymerized fibrin and other factors related to disease-associated coagulopathy, and that it was important to perform additional washes and filtration post-thaw to maximize sample recovery and ensure optimal data quality.

Major immune cell types were defined by manual gating, and the relative frequency of each cell type was evaluated to identify cell types that showed differential abundance between COVID-19 patients and healthy controls. Since all samples were stained using a fixed starting blood volume of 270uL, we first evaluated the absolute cell counts of major immune cell types recovered for each sample. Consistent with other reports, we observed a significant reduction in overall lymphocytes in the COVID-19 patients, most notably in CD8 T cells and NK cells (Figure 6A)^12,13^. When evaluating changes in overall immune cell frequencies within the non-granulocyte PBMC fraction at more granular resolution, we found that COVID-19 patients clearly clustered separately from healthy controls, however there was significant heterogeneity amongst COVID-19 patients (Figure 6B). Some of the consistent features seen across the COVID-19 patients was an increased frequency of plasmablasts, including IgM+ (11-fold average increase, FDR 0.03), IgA+ (8-fold average increase, FDR 0.02) and IgG+ (29-fold average increase FDR 0.02) subsets, CD14+ monocytes (1.3-fold average increase, FDR 0.03), and CD71^hi^ HLADR^hi^ activated CD8 T cells (3-fold average increase, FDR 0.04). In addition, the COVID-19 patients showed a significantly reduced frequency of γδ T cells (2.6-fold average decrease, FDR 0.02) and CD161hi MAIT-like CD8 T cells (5-fold average decrease, FDR 0.04. While the relative frequencies of these populations varied consistently and significantly between the COVID-19 and control groups, it is important to note that the magnitude of the changes varied considerably between patients, consistent with other reports^14^. Several other immune cell subsets including plasmacytoid DCs and other subsets of CD8 T cells and NK cells showed generally decreased frequencies in COVID-19 patients, however, these did not reach statistical significance in this small cohort. In addition to these changes in cell frequency that were consistently seen across most of hospitalized COVID-19 patients, we also noted some immune changes that were specific to certain patients or groups of patients. For example, we noted an increased frequency of CD71^hi^ ICOS^hi^ activated CD4 T cells only in some patients (Fig 6B), suggesting differences in the magnitude of the adaptive T cell response to infection. Evaluation of these heterogeneous responses in relation to other immunological and clinical features of disease severity and outcome is currently ongoing in a larger cohort of hospitalized COVID-19 patients.

**Figure 6.**
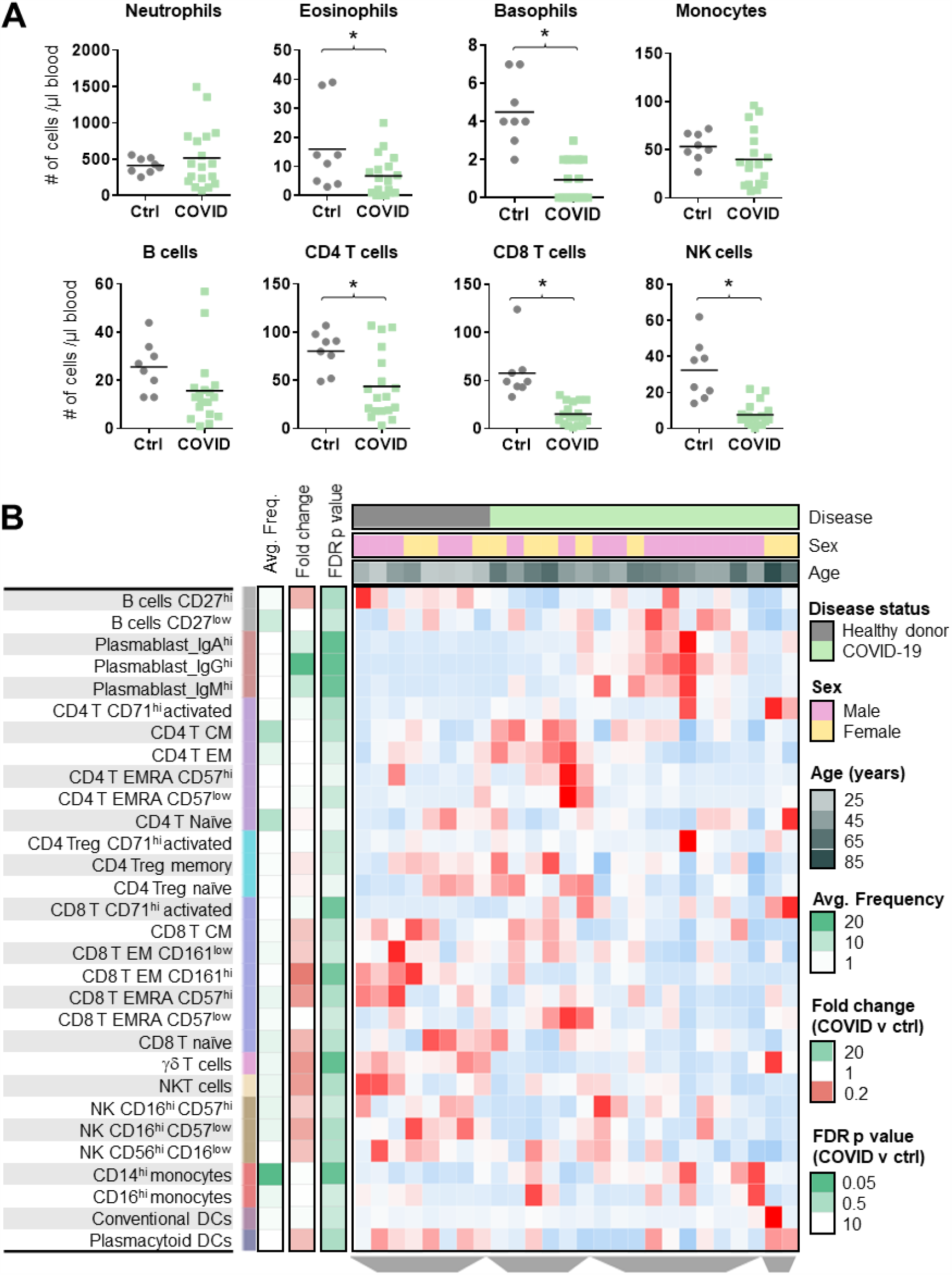
Application of the SmartTube MDIPA workflow to immune monitoring of COVID-19 patients. Aliquots of whole blood from 8 healthy donors and 18 hospitalized COVID-19 patients were stained using the MDIPA-SmartTube workflow and analyzed as described above. (A) Cell counts of major granulocyte, monocyte and lymphocyte subsets COVID-19 patients and controls. (B) Heatmap of relative (z-scored) frequencies for each manually gated subset as a % of PBMCs, summarized for all patients and controls. Each column represents an individual, and column categories indicate whether the subject was a COVID-19 patient or control as well as sex and age. Row categories indicate the average absolute frequency of each gated cell subset, the average fold-change between COVID-19 patients and controls, and the FDR-corrected p-value.

In addition to changes in overall cell type frequencies, we also observed profound changes in the phenotype of circulating monocytes in COVID-19 patients (Figure 7), which were generally more robust correlates of COVID-19 disease status than changes in subset frequency. For example, while the overall frequency of CD14+ monocytes was not consistently altered in the COVID-19 patient cohort, we observed a significant decrease in median HLA-DR expression on monocytes in COVID-19 patients (Figure 7A), consistent with previous reports^15,16^. We also observed a dramatic increase in median CD61 expression on monocytes in all the COVID-19 patients in our cohort, a hallmark of monocyte-platelet aggregates (Figure 7B). Notably, monocyte-platelet aggregates have been shown to be elevated in acute coronary syndromes^17^ and induced *in vitro* models of thrombo-inflammation^10^, and have also been associated with increased risk of mortality in septic patient^18^. Measurement of these complexes may therefore offer an immune correlate of thrombo-inflammation and coagulopathy in COVID-19 patients. While reduced HLA-DR expression and elevated CD61 expression were observed in all COVID-19 patients, we also observed a striking increase in CD169 (sialoadhesin) expression in only a subset of patients (Figure 7C). CD169 is constitutively expressed on subsets of tissue resident macrophages^19^ and is induced on circulating monocytes by type I interferon signaling^20^. Elevated CD169 expression has been reported on circulating monocytes in patients with systemic sclerosis^20^, acute HIV infection^21,22^ and acute zika-virus infection^23^. Expression of CD169 on monocytes in COVID-19 patients may therefore offer a cellular biomarker of acute type I IFN signaling. It is intriguing to consider whether the considerable heterogeneity in CD169 expression amongst our cohort of COVID-19 patients may reflect constitutive differences in anti-viral IFN responses between patients^24^ or differences in disease course, which are ongoing topics of investigation in this cohort.

**Figure 7.**
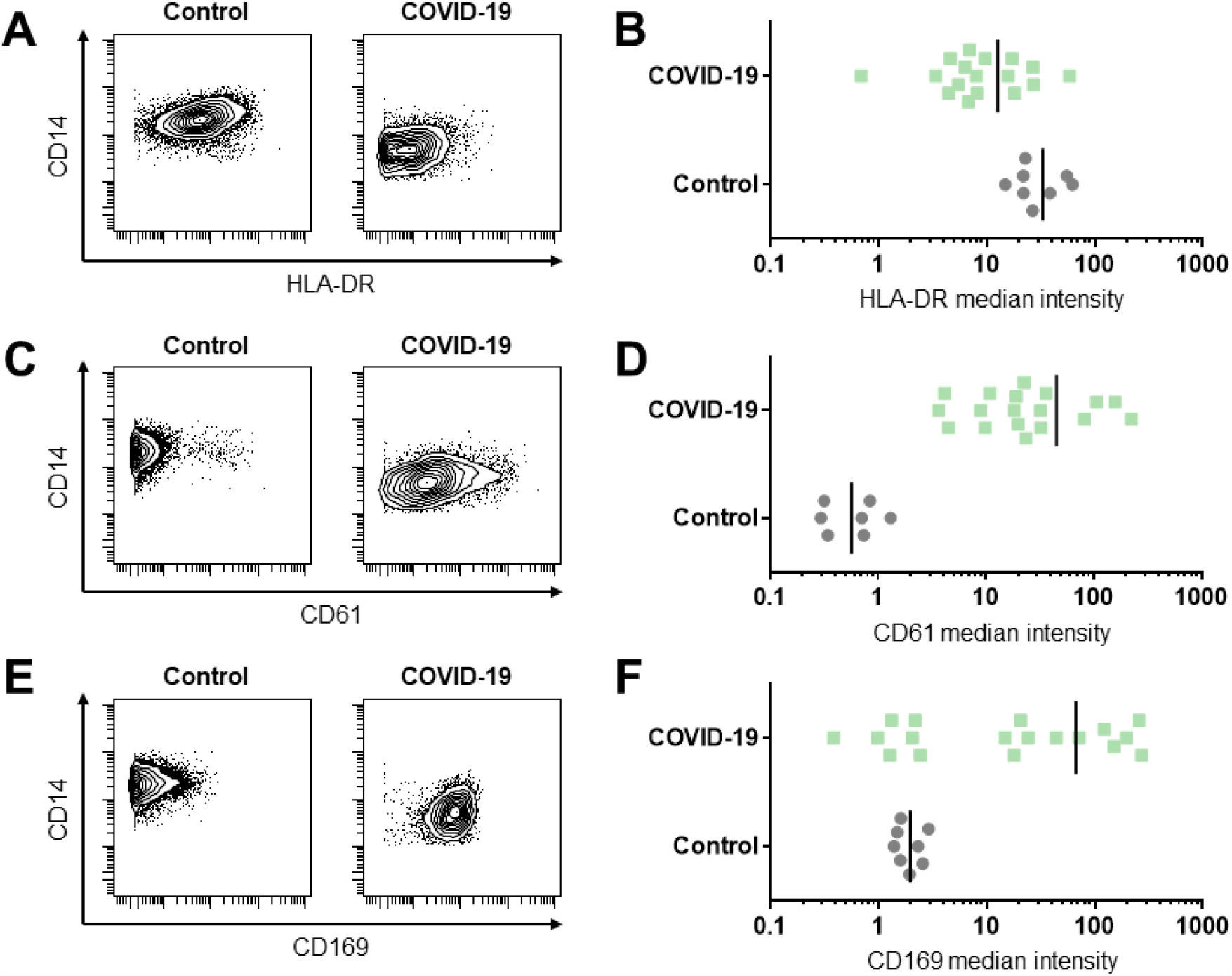
Alterations in circulating monocyte phenotype in COVID-19 patients. Aliquots of whole blood from 8 healthy donors and 18 hospitalized COVID-19 patients were stained using the MDIPA-SmartTube workflow and analyzed as described in Figure 6 and CD14+ monocytes were manually gated. (A) Biaxial plots from a representative patient and control and (B) median expression across all patients and controls of HLA-DR. (C) Biaxial plots from a representative patient and control and (D) median expression across all patients and controls of CD61. (E) Biaxial plots from a representative patient and control and (F) median expression across all patients and controls of CD169.

## Conclusions

Overall, we believe that the data provided here provide a validation of a streamlined, sample-sparing whole blood immune monitoring workflow (Figure 1) and offer data to support important considerations in terms of vacutainer selection, storage duration prior to staining, and the incorporation of additional markers to supplement the core MDIPA panel. We also demonstrate and discuss important considerations in the successful application of this protocol to characterize immune responses in COVID-19 patients. Comprehensive immune profiling using this assay identifies overall lymphopenia in COVID-19 patients, but also highlights acute adaptive immune responses in some patients, represented by elevated frequencies of plasmablasts and activated CD8 T cells, and also profound changes in myeloid cell phenotype, including reduced HLA-DR expression, elevated CD61 and CD169 expression. While all COVID-19 patients in this cohort showed some level of immune changes relative to healthy controls, we observed considerably cellular heterogeneity among patients, and studies are ongoing to relate this heterogeneity to relevant clinical metrics of disease severity and progression in a larger cohort.

While this protocol already offers a considerably streamlined workflow that requires only 2 pipetting steps, this still requires an appropriate BSL2-approved environment to process the blood samples, and the ability to accurately pipette and time the addition of the necessary volumes, which may still not be possible at some resource-limited clinical collection sites. Thus, a next logical step towards enabling even broader adoption of these workflows would be to pre-package the lyophilized antibody panel as part of a syringe based fixed-volume blood collection device, such as TruCulture system [18], and to automate the timing and addition of the stabilization buffer and cryopreservation steps, using a device such as the SmartTube base station.

## Data Availability

Annotated FCS files have been deposited in FLOWRepository under IDs: FR-FCM-Z2XA and FR-FCM-Z2XB

## Acknowledgments

We thank all the members of the Mt. Sinai Human Immune Monitoring Center and the many individuals who contributed to the Mt. Sinai Precision Immunology COVID-19 Repository effort. This work was supported by NIH U19AI118610 (M.M. and A.R.), U24AI118644 (M.M. and A.R., U24CA224319 (S.G. and A.R.). Helios mass cytometry instrumentation at the Human Immune Monitoring Center was obtained with support from S10OD023547.

## Data availability

Annotated FCS files have been deposited in FLOWRepository under IDs: FR-FCM-Z2XA and FR-FCM-Z2XB

## Supplementary Material

**Supplementary Figure S1.**
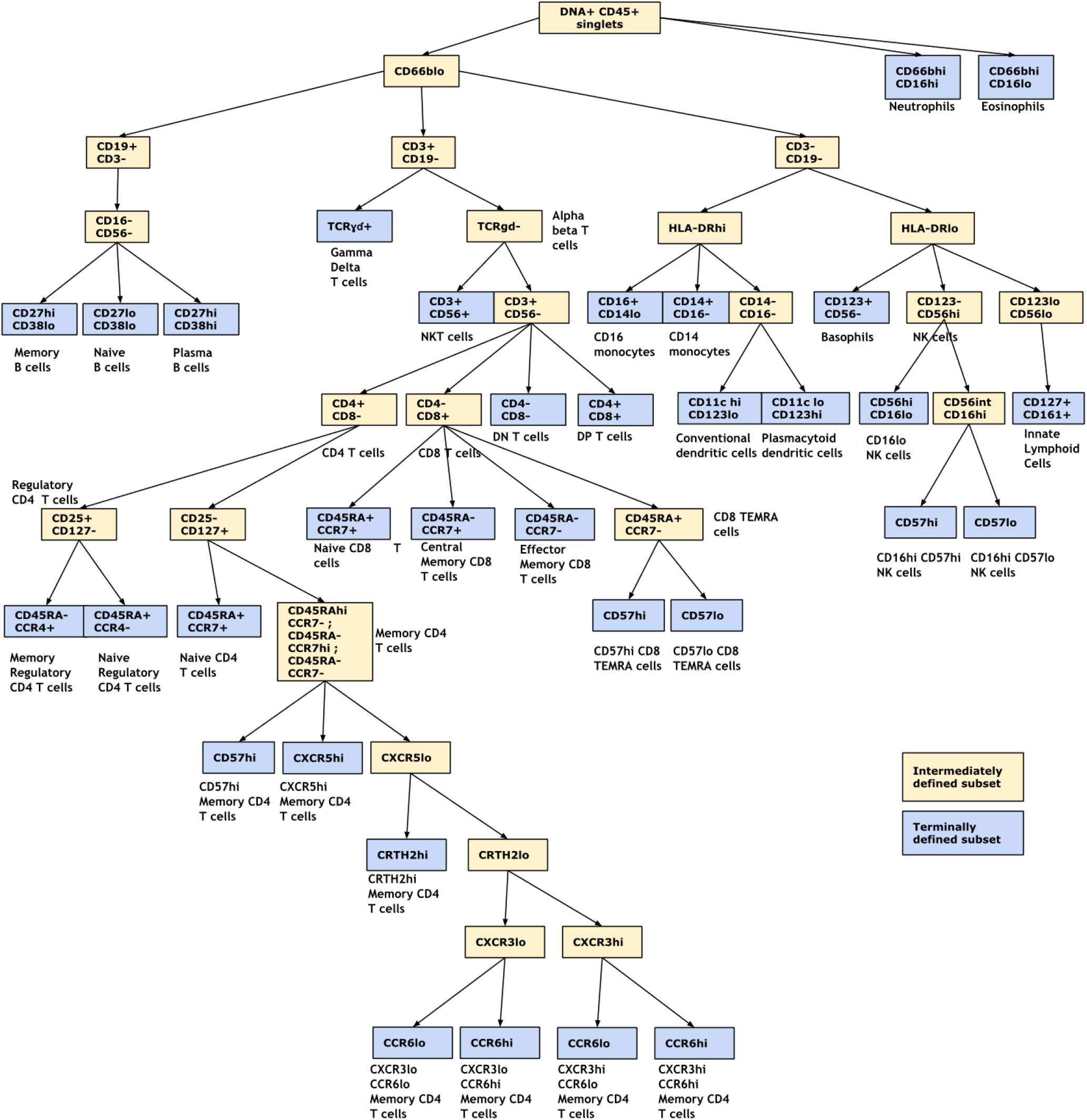
Schematic of the manual gating hierarchy used to define immune populations with the MDIPA panel.

**Supplementary Figure S2.**
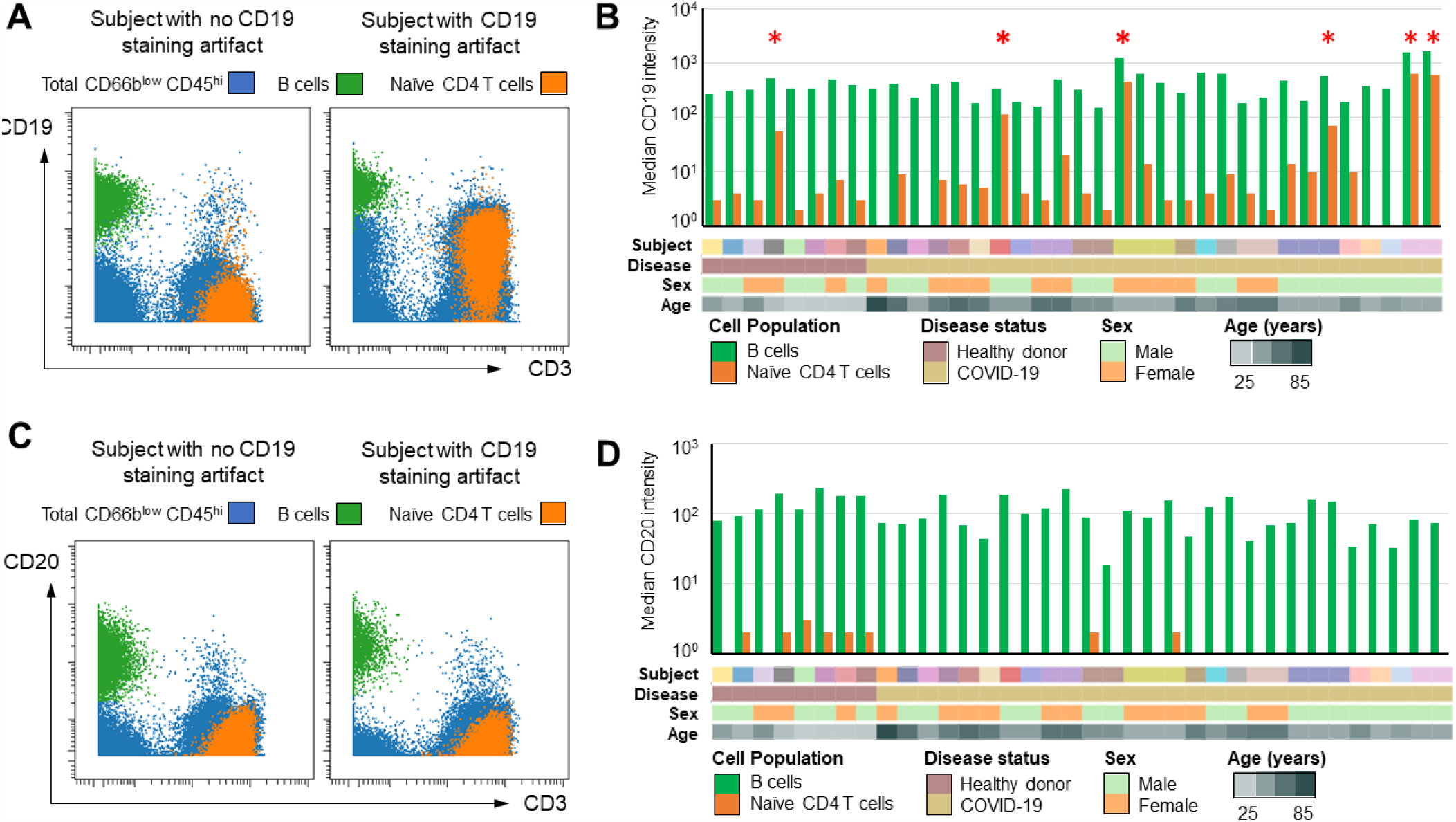
An artifact of aberrant CD19 staining on non-B cells in some individuals when using the MDIPA panel. Whole blood samples from 28 individuals were stained and analyzed with the MDIPA-SmartTube workflow as described. (A) Representative biaxial plots showing CD19 vs CD3 staining in blood from a non-affected donor (left) and an affected donor (right). (B) Summary statistics across all 28 individuals showing the median expression of CD19 on paired B cells and naïve CD4 T cells. Red asterisks highlight individuals showing particularly high levels of non-specific CD19 staining. Column category color bars indicate samples collected from the same subject, together with COVID-19 disease status, sex and age. (C) Corresponding biaxial plots showing CD20 vs CD3 staining from the same individuals shown in (A). (D) Corresponding summary statistics of median CD20 expression on paired B cells and naïve CD4 T cells for the same individuals shown in (B).

**Supplementary Figure S3.**
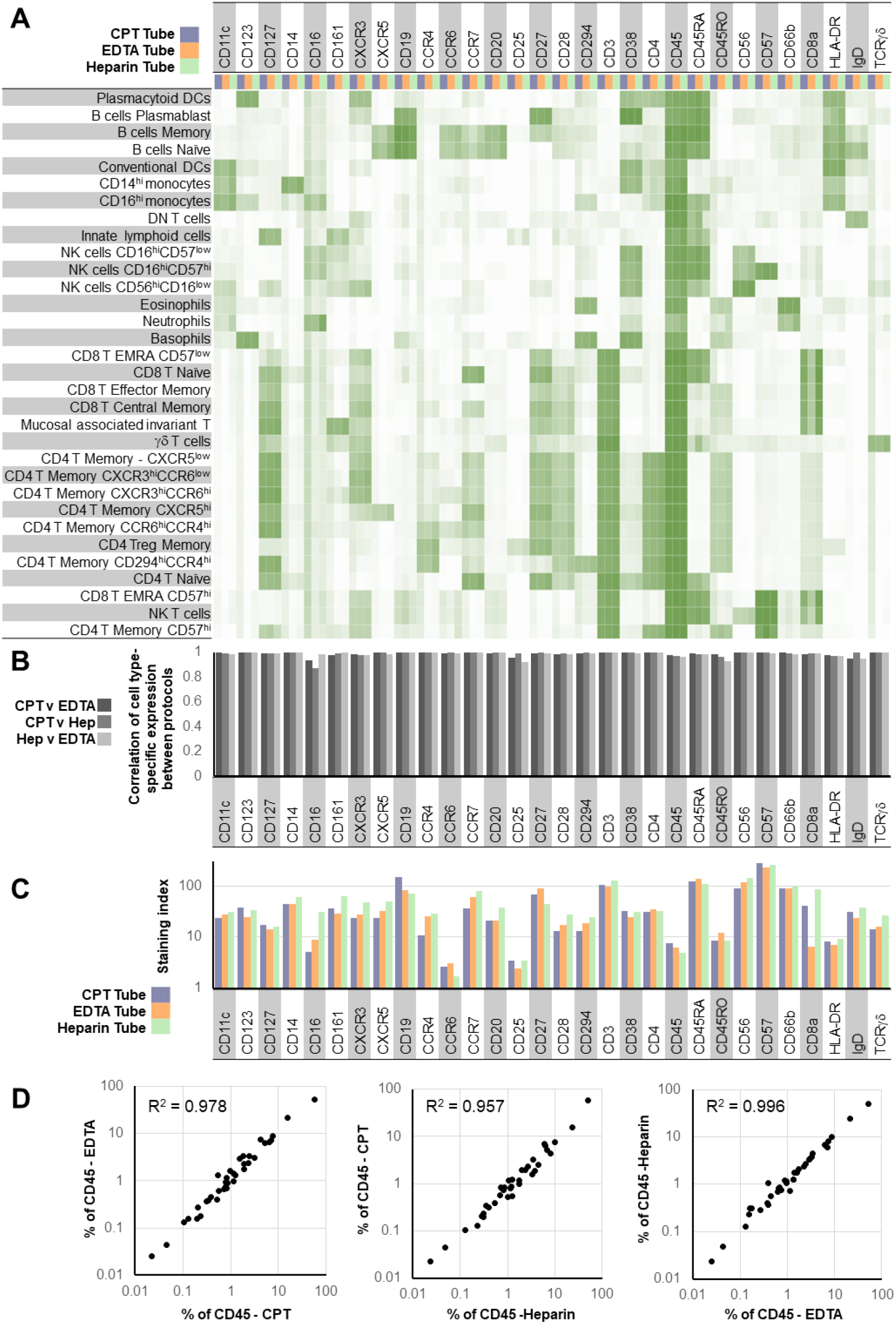
The SmartTube-MDIPA workflow can be successfully applied to whole blood collected using multiple kinds of anticoagulant tubes. Whole blood was collected from the same healthy donor into a sodium heparin tube, an EDTA tube or a CPT-citrate tube. Aliquots from each tube were stained with the MDIPA-SmartTube workflow. (A) Heatmap of median marker expression of each marker on manually gated immune subsets in blood collected in each of the three tube types. (B) Pearson’s coefficient of the correlation of the expression of each marker across all subsets in each of the three tube types. (C) Staining index of each marker in each of the three tube types. (D) Correlation of overall population frequencies for each gated cell type between the three tube types.

**Supplementary Figure S4.**
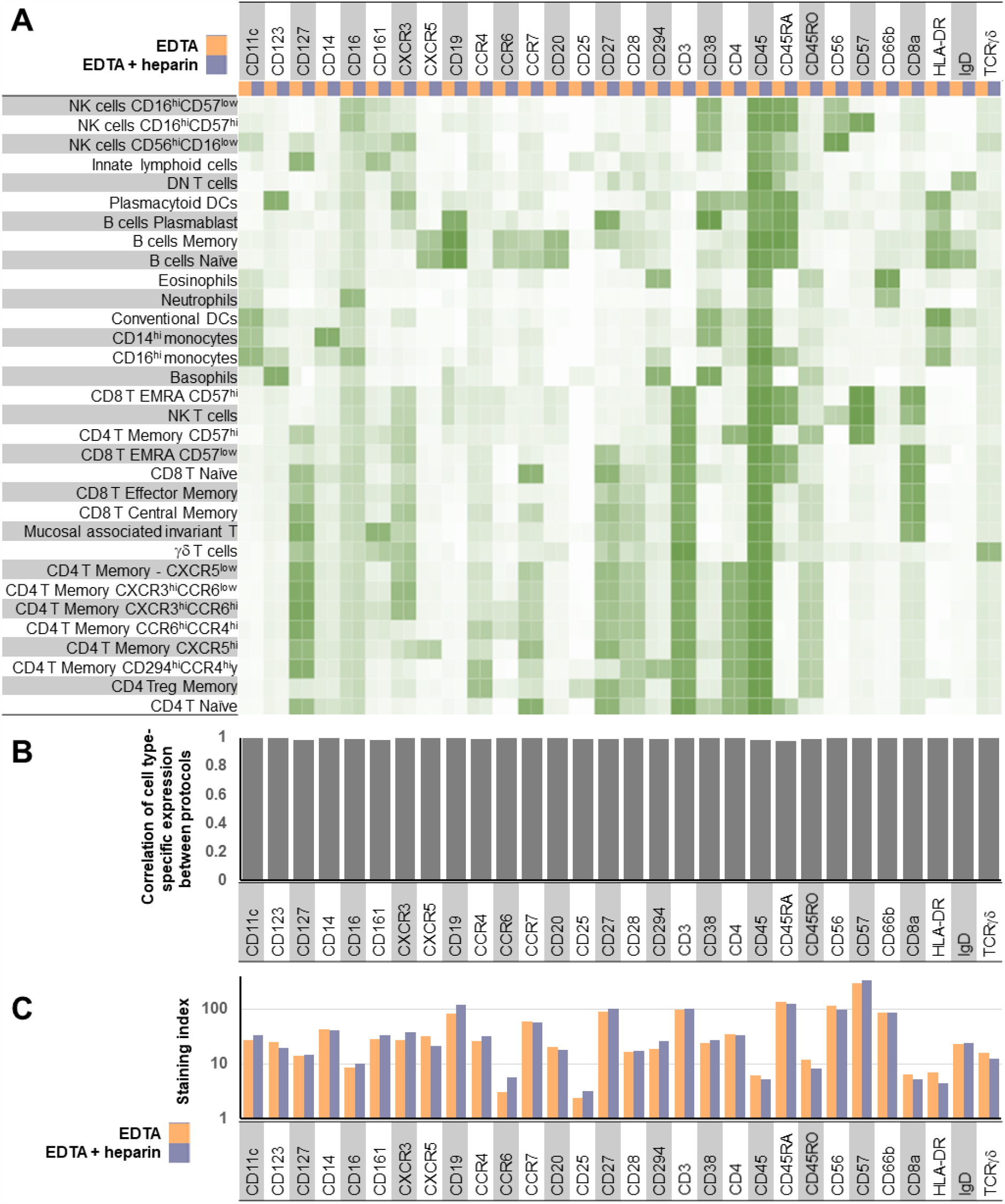
Additional heparin supplementation has no appreciable impact on staining quality or non-specific background staining of blood collected in EDTA tubes. Parallel aliquots of healthy donor whole blood collected in an EDTA tube were stained with the MDIPA-SmartTube workflow with or without additional heparin supplementation. (A) Heatmap of median marker expression of each marker on manually gated immune subsets using the two workflows. (B) Pearson’s coefficient of the correlation of the expression of each marker across all subsets using the two workflows. (C) Staining index of each marker using the two workflows.

**Supplementary Figure S5.**
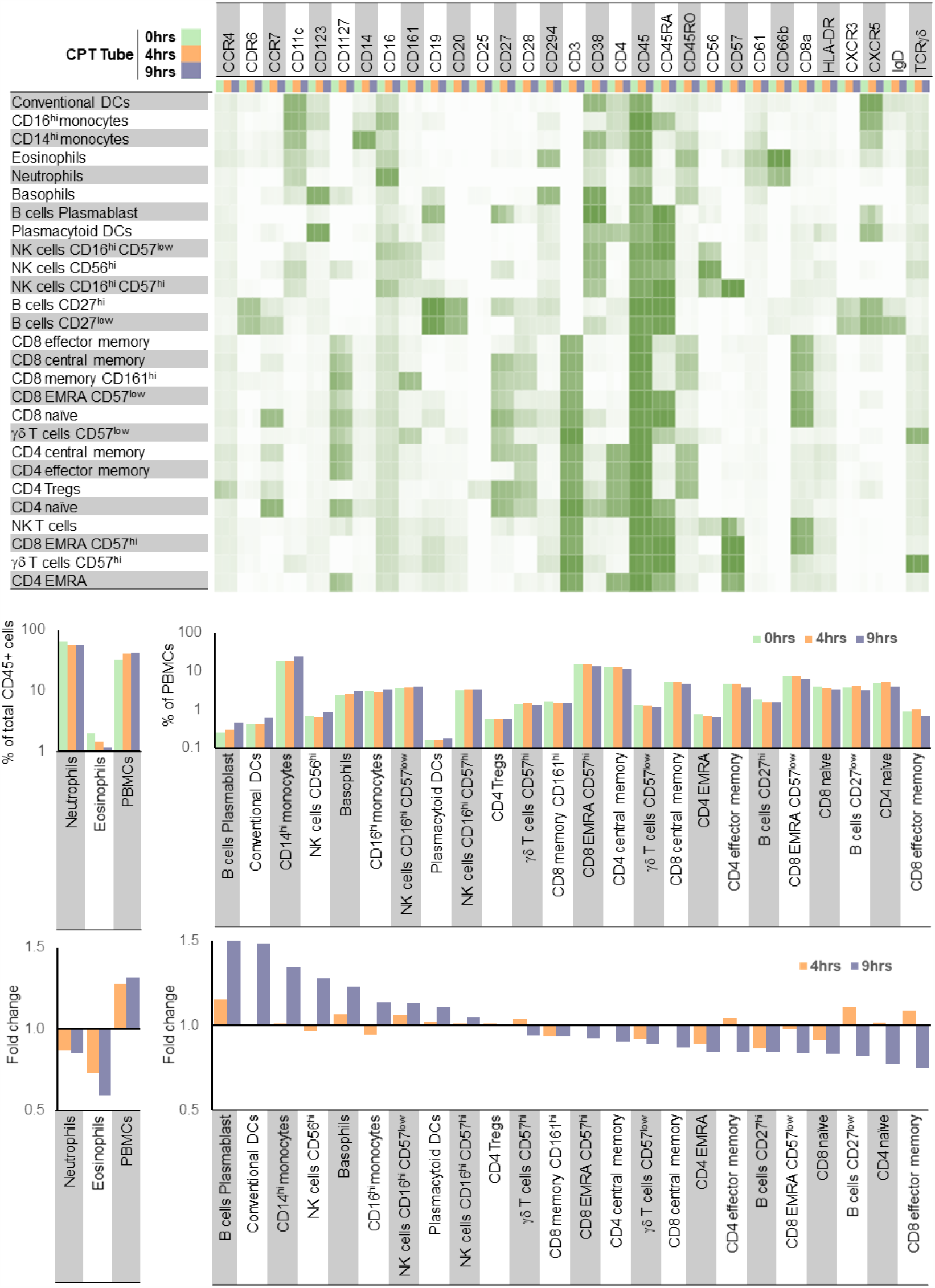
Impact of delayed processing time on staining patterns and population frequencies. Blood was collected from a healthy donor in a CPT-Citrate tube and left resting on a bench at room temperature with no agitation to replicate delayed processing after collection. At time intervals of 0, 4 and 9hrs after collection, aliquots of blood were removed from the tube and stained with the MDIPA-SmartTube workflow. After thawing, the samples were also stained with a supplemental CD61 antibody to evaluate potential formation of platelet-leukocyte aggregates. (A) Heatmap of median marker expression of each marker on manually gated immune subsets at each of the timepoints. (B) Frequency of each gated immune cell subset at each timepoint. (C) Fold change in the frequency of each subset at 4hrs and 9hrs relative to time 0.

